# Tensions between research and public health: modelling the risks and benefits of SARS-CoV-2 vaccine field trials versus human infection challenge studies

**DOI:** 10.1101/2020.05.18.20106187

**Authors:** George S Heriot, Euzebiusz Jamrozik, Michael J Selgelid

## Abstract

**Background:** Human infection challenge studies (HICS) with SARS-CoV-2 are under consideration as a way of accelerating vaccine development. We evaluate potential vaccine research strategies under a range of epidemic conditions determined, in part, by the intensity of public health interventions.

**Methods:** We constructed a compartmental epidemiological model incorporating public health interventions, vaccine efficacy trials and a post-trial population vaccination campaign. The model was used to estimate the duration and benefits of large-scale field trials in comparison with HICS accompanied by an expanded safety trial, and to assess the marginal risk faced by HICS participants.

**Results:** Field trials may demonstrate vaccine efficacy more rapidly than a HICS strategy under epidemic conditions consistent with moderate mitigation policies. A HICS strategy is the only feasible option for testing vaccine efficacy under epidemic suppression, and maximises the benefits of post-trial vaccination. Less successful or absent mitigation results in minimal or no benefit from post-trial vaccination, irrespective of trial design.

**Conclusions:** SARS-CoV-2 HICS are the optimal method of vaccine testing for populations maintained under epidemic suppression, where vaccination offers the greatest benefits to the local population.

## INTRODUCTION

Public health interventions have successfully limited the initial rapid transmission of coronavirus disease 2019 (COVID-19) in many countries, but the relaxation of these measures without achieving elimination or high levels of herd immunity risks epidemic recrudescence. Over 100 SARS-CoV-2 vaccines are currently under development[1] but there are controversies regarding the appropriate method(s) for testing the efficacy of these candidates prior to widespread public health use.

Vaccine field trials depend on natural infection of individuals in the population and are therefore most feasible during periods of rapid transmission, which public health measures have sought to mitigate and/or prevent. One consequence of a high transmission epidemic (during which a field trial might be conducted) is a high level of herd immunity, which erodes the benefit of (post-trial) vaccination to the surviving population. The goals of public health measures and those of SARS-CoV-2 vaccine research are therefore intimately related and sometimes in conflict.

Human infection challenge studies (HICS) involve the intentional infection of research participants and are conducted with many pathogens including pandemic influenza.[2] Challenge studies have been proposed as a method of accelerating SARS-CoV-2 vaccine testing,[3, 4] because they typically involve far fewer participants and require less time to conduct than field trials.[5, 6] Although field trials usually take much longer to demonstrate vaccine efficacy, the rapid transmission of COVID-19 has led to suggestions that HICS may not in fact accelerate vaccine development relative to field trials. [7]

In this article, we use an epidemiological compartment model to illustrate the effects of different epidemic conditions on the duration and local benefits of vaccine efficacy trials and the marginal risks to HICS participants. This permits an evaluation of optimal vaccine research strategies under different public health policies of non-vaccination transmission control measures ranging from moderate measures aimed at mitigation to strict measures aiming for suppression.

## MATERIALS AND METHODS

### Compartmental epidemiological model

We constructed a standard non-age-structured compartmental epidemiological model (Figure A1, Supplementary Methods) without vital dynamics describing the number of individuals in a fixed population who are *Susceptible* to infection (*S*), *Exposed* (incubating but not yet infectious, *E*), *Infected* (and able to transmit infection, *I*), *Removed* (neither infectious or able to be infected, *R*), and *Vaccinated* (but not yet immune, *V*). This simple deterministic model has a number of structural assumptions including homogenous mixing of a closed population, no stratification of transmissibility by subpopulations, and complete and permanent immunity after natural infection. The flow of individuals between compartments is governed by a set of ordinary differential equations presented in the Supplementary Methods.

The model starts on day 1, has a period of one day, and runs to a horizon of ten years after the initial introduction of transmission control measures.

### Disease characteristics

The disease characteristics used in this exploration are taken from current descriptions of COVID-19 (Table 1). There is still substantial uncertainty around these estimates and how they apply to a given setting, and there are insufficient data on which to base credible parameter distributions. Our estimates reflect those being used to guide Australia’s public health response.[8]

**Table 1.** Disease characteristics.

| Parameter | Modelled value |
| --- | --- |
| Basic reproductive number ($R_0$) | 2.53[8] |
| Latent period prior to infectivity | 3.2 days[8] |
| Duration of infectivity | 9.68 days[8] |
| Time from infection to death | 22 days[29] |
| Population infection fatality ratio (IFR) | 0.66%[29] |
| IFR among HICS participants | 0.03%[29] |
| HICS human infection challenge study |  |

### Setting

The model was run on a stable closed population the size of the Australian state of Victoria.[9] On day one of the model, the population is entirely (and uniformly) susceptible to infection except for a single exposed individual incubating infection. Transmission control measures are applied on the first day where the expected cumulative number of infection-related deaths rises to one (calculated to be day 77 on the basis of the parameters described so far, and the end of March 2020 in real-world Victoria). At the introduction of these measures, the cumulative prevalence of infection in the model population (*E*+*I*+*R*) remains very low (0.02%).

The effective reproductive number achieved during the first period of transmission control is 0.5, which is the estimated figure achieved in Victoria during the current phase of restrictions.[10] After 42 days of initial restrictions, the effective reproductive number rises (to R_eff_) as a result of the partial relaxation of transmission control measures, as is currently planned.[11] R_eff_ is modelled as a continuous variable ranging between 0.8 (achieving eradication in the study population by three years) and 2.5 (an unmitigated epidemic). The effective reproductive number achieves stability over seven days (one infectious period) after each change via a seven-day moving average.

### Trials of a candidate vaccine

We assumed the first candidate vaccine has efficacy for the trial outcome, noting that this assumption removes a key benefit of HICS (the parallel comparison of multiple candidate vaccines)[12]. Vaccine efficacy (the complement of the relative risk reduction for infection) was considered as a variable in the model with values of 50%, 70% and 90% (derived from WHO specifications[13]). We assumed that the vaccine does not alter the course of subsequent infection either through vaccine-enhanced disease or partial protective immunity. In both the field trial and HICS, participants receive two doses of vaccine 28 days apart. We assumed a mean time to protective immunity after the first dose of vaccine to be 14 days, implying 86% of maximal vaccine efficacy is achieved by the time of the second dose and 98% by 28 days after the second dose.

Both efficacy trials commence on day 260, six months after the initial imposition of transmission control measures, when the candidate vaccine supported by preclinical data becomes available in sufficient quantity.[14] Both trials were specified as 1:1 randomised placebo-controlled trials examining the dichotomous outcome of SARS-CoV-2 infection demonstrated by a perfect test. This outcome is much more feasible in the setting of a HICS than a large field trial, but was chosen in the absence of reliable clinical or laboratory surrogate markers of infection.[15-17] The trials are designed to have a 90% power to detect vaccine efficacy above the minimum desirable vaccine efficacy (set at 30%)[18] with a one-sided alpha error of 2.5% using a frequentist approach.

The group size in the field trial is set to 10,000, roughly the size of the largest field trials of novel vaccines in recent years.[19-21] Infections occurring during the first 56 days after the first dose of vaccine are censored, and participants infected within the censoring period are excluded from the primary per protocol analysis (see Power calculations in the Supplementary Methods). Participants are assumed to be independent with regards to outcome. The field trial ends seven days after sufficient exposures have occurred to provide 90% statistical power for the vaccine efficacy under consideration. This duration assumes either perfect prediction or intensive independent monitoring[18] of both the rate of incident infection and vaccine efficacy during trial design, and does not include a statistical safety margin. In order to collect short-term safety data, the minimum duration of the trial is set to 70 days (42 days after the second dose of vaccine).[4] As the vaccinated trial participants remain in the community, the vaccination event is included in the model as a one-off transfer from the *S* to the *V* compartments.

HICS participants are inoculated with challenge strain 28 days after the second dose of vaccine. We have not modelled sequential inoculation cohorts as a precaution against vaccine enhanced disease,[22] as the counterfactual large field trial presupposes that this risk would be considered minimal after preceding phases of vaccine development. The HICS ends seven days after challenge, when all incident infections are identifiable. The HICS participants are assumed to be isolated until they are *Removed* and do not contribute to the number of *Exposed* individuals in the population. The HICS is followed one week later by an expanded safety trial where 5,000 participants are vaccinated and monitored for 70 days for safety and immunogenicity outcomes (and move from the S to the V compartments).[22]

### Mass vaccination program

We modelled an indiscriminate mass vaccination program (as would be expected in the absence of a reliable and scalable method of determining either pre-existing immunity or active or incubating infection) where 1% of the population receives their first dose of vaccine each day for a total of 100 days. Vaccination of the model population commences 90 days after the completion of the vaccine efficacy trial, by which time more than 60,000 doses need to be available each day for administration to the model population. Both of these assumptions are highly optimistic regarding both the rapidity and reach of the campaign. The indiscriminate nature of vaccine delivery allows the vaccination rate among *Susceptible* individuals to match the population rate.

Transmission prevention measures are lifted immediately after the last day of the vaccination campaign, or at three years after imposition, whichever is earlier. The effective reproductive number from this point is set to 2.2, reflecting long-term behavioural changes.[23]

A timeline of the model conditions is presented in Figure A2 in the Supplementary methods.

### Outcomes

The model outcomes of interest were the ten-year cumulative incidence of COVID-19 mortality in the population; the duration of a vaccine field trial conducted in this population; the mortality benefit to the population of a vaccination campaign following either of the two trial methods compared to no vaccine being available; and the absolute mortality risk to HICS participants compared to the background risk of infection. We specifically consider two idealised public health policies: i) optimal “mitigation”, where R_eff_ is set to achieve the lowest cumulative population mortality in the absence of a vaccine; and ii) “suppression”, where R_eff_ is reduced below 1.0 by non-vaccination transmission control measures.

All primary analyses were conducted as two-way sensitivity analyses varying both R_eff_ and vaccine efficacy across the values described above. Additional sensitivity analyses examining the effect of a delay to candidate vaccine availability and higher population prevalence of infection prior to the introduction of transmission control measures are presented in the Supplementary Results.

## RESULTS

### Cumulative population mortality under epidemic mitigation policies

In a low-prevalence population, transmission control measures without a vaccine result in a cumulative population mortality from COVID-19 between 0.3% and 0.54% over a ten-year period, corresponding to a cumulative prevalence of infection between 45% and 82% (Figure 1). Optimal mitigation (R_eff_ 1.41) achieves the lowest cumulative population mortality (0.3% without a vaccine) and a peak mortality rate of 2.1 deaths per 100 000 population per day manageable with current Victorian physical critical care capacity.[24, 25]

**Figure 1.**
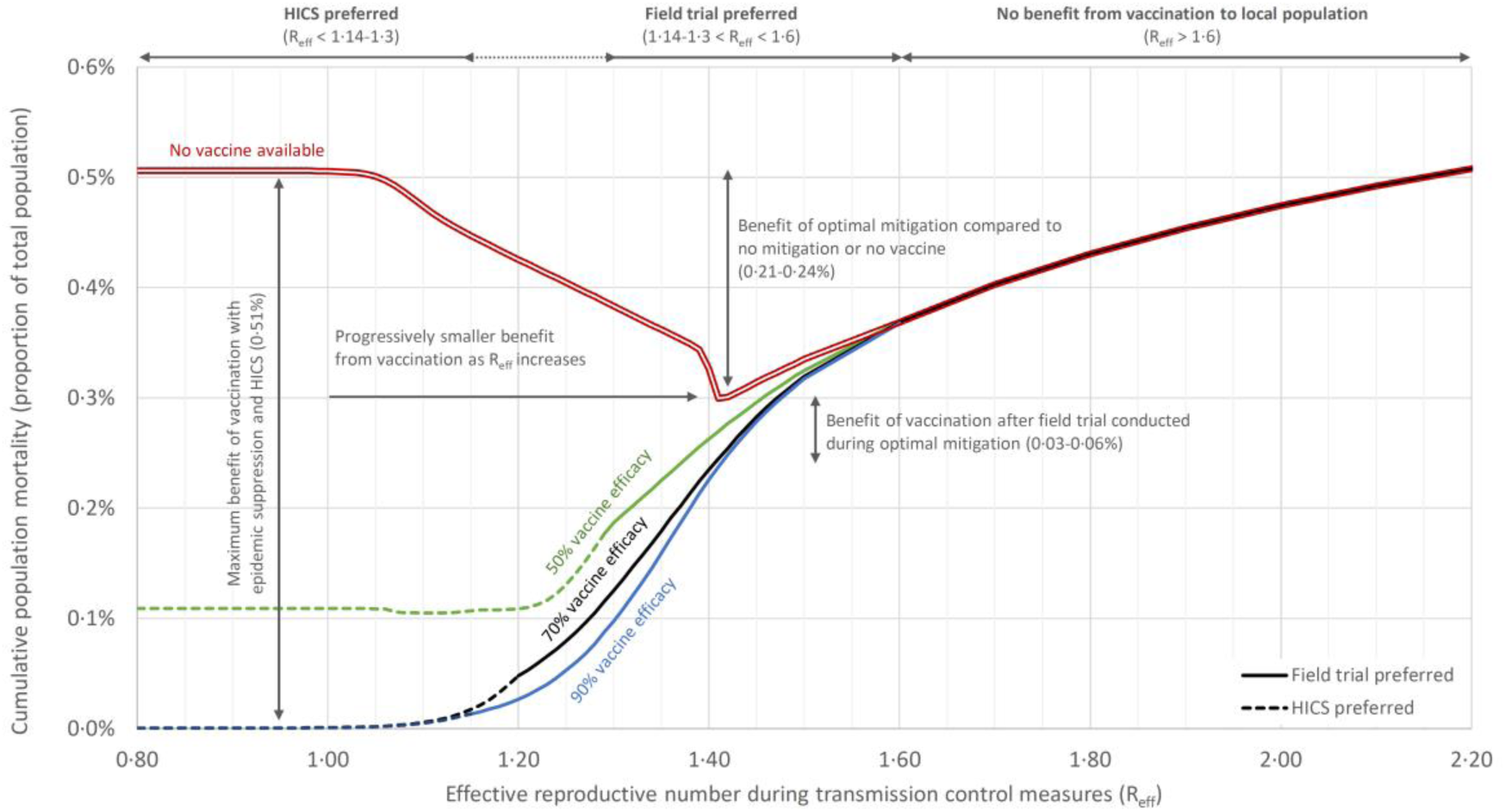
Population mortality benefit of vaccination after trials of vaccine efficacy. The maximum benefit of vaccination follows epidemic suppression and a HICS. Field trials can offer similar benefits (with longer trial durations) at low vales of R_eff_, but fail to report when the effective reproductive number falls below 1.03 to 1.08. Lifting of suppressive transmission control measures where no vaccine is available results in an unmitigated epidemic, thus no long-term population mortality benefit. Neither trial type offers benefit to the population with reproductive numbers above 1.6. Mitigation alone offers substantial benefit even without considering excess mortality from overwhelmed healthcare resources.

### Minimal benefit of vaccine programs after suboptimal epidemic mitigation

In the absence of effective mitigation (R_eff_ greater than 1.6; Figure 1) vaccination does not substantially accelerate epidemic termination above immunity derived from natural infection. A vaccination campaign conducted after efficacy testing during the epidemic (whether via HICS or field trial) provides little if any population benefit because of high post-epidemic herd immunity.

### Vaccine efficacy trials under epidemic mitigation policies

A vaccine field trial conducted under optimal mitigation (R_eff_ 1.41) and commencing six months after the imposition of transmission control measures would take between 70 and 92 days to achieve 90% power, resulting in approximately 15 months of transmission control measures (Figure 2). A vaccination campaign following this demonstration of efficacy would reduce the cumulative population mortality by 0.03-0.06% below the effect of transmission control measures alone. This benefit is highly sensitive to additional delays in the vaccine development timeline (Figures A3 and A4, Supplementary Methods). In contrast, a HICS commencing at the same time followed by an expanded safety trial requires 140 days and post-trial vaccination reduces the cumulative population mortality to a lesser degree (0.01%).

**Figure 2.**
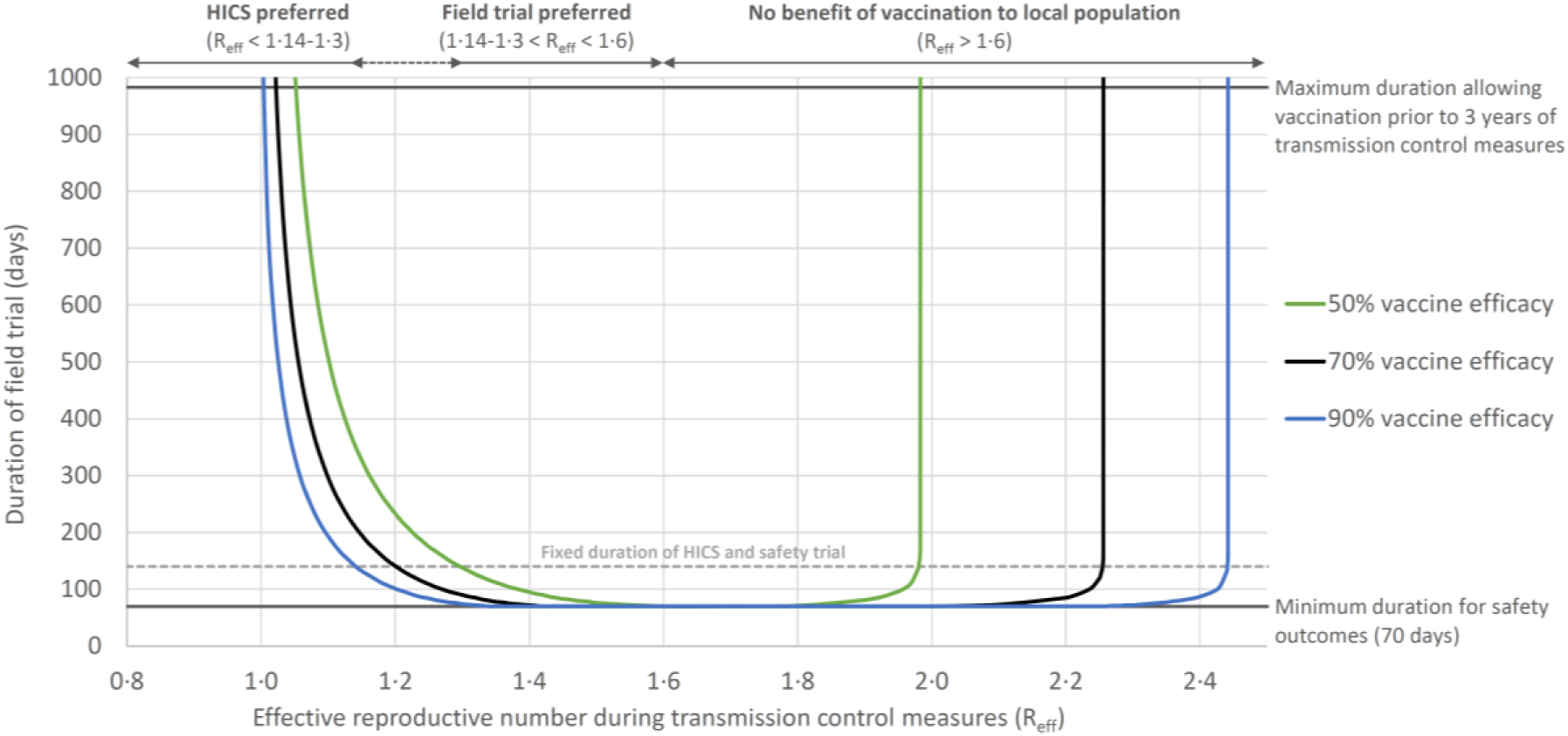
Duration of vaccine field trial by intensity of transmission control measures and vaccine efficacy. The modelled field trial fails to achieve 90% statistical power at both low and high values of R_eff_ due to an insufficient event rate; at low values, due to strict transmission control polices; at high values, due to herd immunity causing natural waning of transmission in the latter phases of the epidemic.

Stricter (sub-suppressive) transmission control measures (e.g., R_eff_ 1.10) result in the substantial prolongation of a field trial of the same size (to between 194 and 508 days, or 19 to 29 months of transmission control measures). A larger proportion of the population would remain susceptible by the time of the subsequent vaccination campaign, which would reduce the cumulative population mortality by up to 0.50% (Figure 1). A HICS and safety trial conducted under these conditions would still report in 140 days and would achieve similar (or marginally greater) reduction in cumulative population mortality to the field trial. The advantage of HICS over field trials in terms of duration and population outcomes persists with values of R_eff_ up to 1.14-1.3 (depending on vaccine efficacy).

### Vaccine efficacy trials under a policy of epidemic suppression

The greatest potential for population benefit from vaccination occurs when epidemic suppression (R_eff_ less than 1.0) is maintained until mass vaccination has been achieved (Figure 1). Under these conditions, widespread use of a vaccine with efficacy greater than the calculated herd immunity threshold (roughly 61%) has the potential to prevent all but a handful of deaths. However, even with 90% vaccine efficacy and 20,000 total participants, a field trial would fail to achieve 90% power under these conditions. In this context, HICS (requiring 24, 70 and 312 participants at vaccine efficacy 90%, 70% and 50%, respectively) are the only mechanism of demonstrating vaccine efficacy within, and achieving maximal benefit for, the local population.

### Excess risk to HICS participants

The excess risk of death among HICS participants due to deliberate inoculation above that from natural infection in the population depends on the background risk of transmission and whether a vaccine would be available to potential participants if they did not participate in the trial. Figure 3 presents the absolute excess mortality risk for HICS participants if a vaccine were available after others volunteered for the study (solid lines), and the excess mortality (or net benefit) should there be no volunteers and thus no vaccine (dashed lines). Even at its maximum modelled value with a pessimistic estimate for the IFR among healthy HICS participants (see Table A1 in the Supplementary Methods), the excess mortality risk to participants remains very small (1.8 per 10,000 or 0.018%).

**Figure 3.**
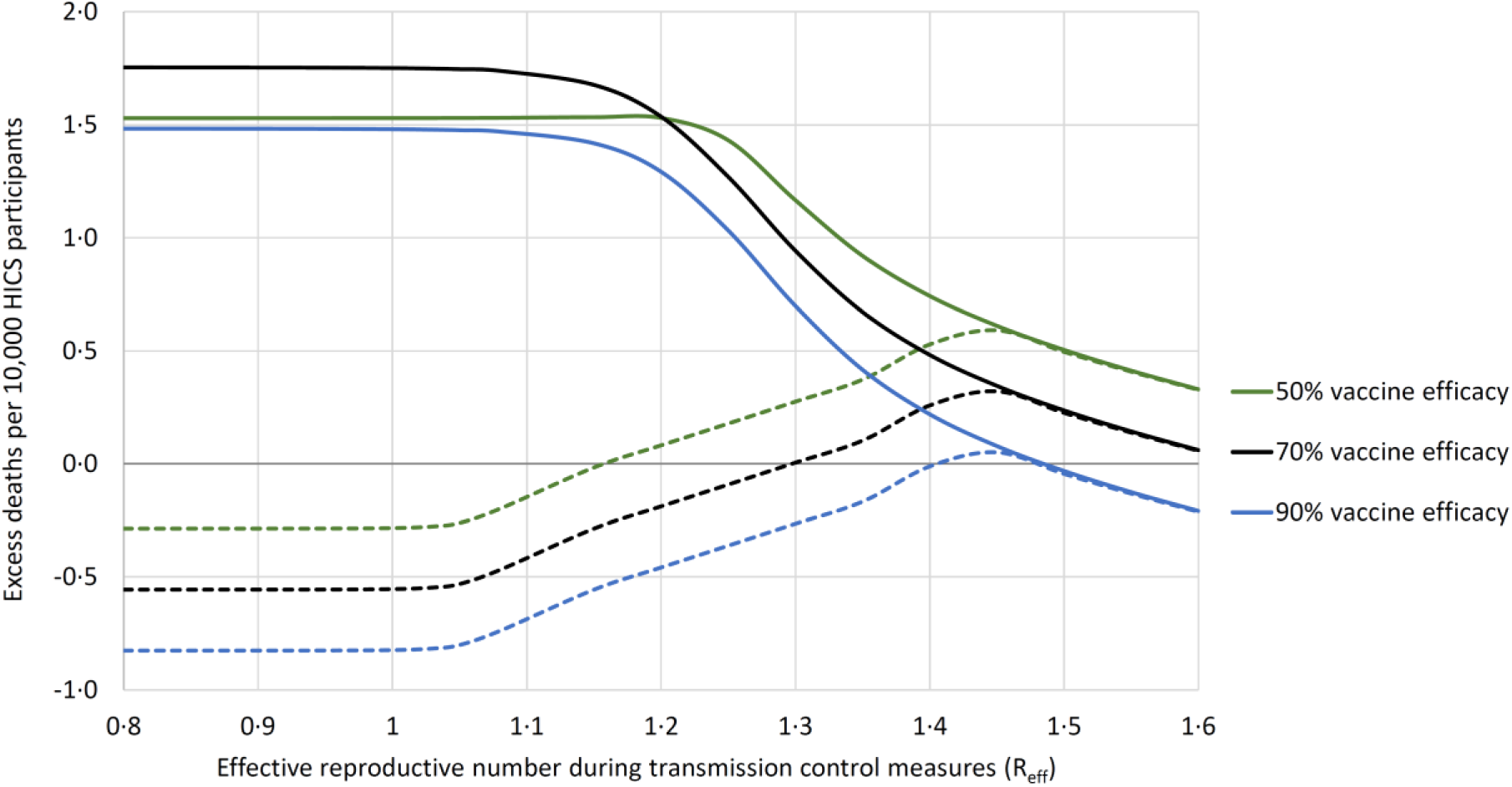
Excess mortality among human infection challenge study (HICS) participants. Solid lines denote risk compared to a vaccine being made available by others volunteering; dashed lines denote net benefit to participants if a vaccine were not otherwise made available. The values of R_eff_ in this figure are limited to those where significant vaccination benefit is available to the population from which HICS participants are drawn. The excess risk with 50% vaccine efficacy is abrogated between R_eff_ 0.8 and 1.2 because protection from vaccination alone is below the herd immunity threshold (roughly 61%) increases the risk to HICS non-participants after lifting of transmission control measures. Similarly, the reduced benefit of vaccination occurring after peak transmission increases the relative benefit of effective vaccination within the HICS at higher values of R_eff_.

## DISCUSSION

Public health measures during a COVID-19 epidemic strongly influence the feasibility of vaccine field trials. Since field trials rely on incident infection to demonstrate vaccine efficacy, there is a natural tension between the goals of transmission control measures and those of field trial research. Public health measures and other epidemic conditions influence the magnitude of potential benefits and risks of SARS-CoV-2 vaccine trials, including the potential benefits of post-trial vaccination campaigns and the marginal risks among HICS participants. Although high background risk of infection results in reduced marginal risks to HICS participants,[12] a substantially greater public health benefit results from HICS conducted where background risk is low during the period before vaccine availability (i.e., in a population opting for epidemic suppression).

### Vaccine field trials under epidemic mitigation

Mitigation policies aim to reduce overall cumulative mortality (and unsustainable healthcare demand) without necessarily relying on the emergence of an effective vaccine, but may also provide conditions conducive to vaccine field trials as a secondary benefit. In the model presented above, transmission control measures consistent with mitigation (R_eff_ 1.1 to 1.6) allow accumulation of sufficient events in a field trial while retaining some benefit from a post-trial vaccination campaign. The overall benefit of post-mitigation vaccination is nevertheless substantially smaller than vaccination of a population maintained in a state of epidemic suppression, and this local benefit deteriorates further with any additional prolongation of the vaccine development timeline (Figure A3, Supplementary Results). Eventually, when transmission slows due to accumulated population immunity derived from infection (or alternative durable transmission control measures), field trials become infeasible (Figure 2, or the 2015-16 Zika virus epidemic[26]).

Major practical caveats of field trials under mitigation policies include (i) reliance on a sophisticated public health response able to calibrate transmission control measures to within the required range for R_eff_ and (ii) progressive challenges of identifying (with imperfectly sensitive serological assays) and recruiting large numbers of susceptible (never-infected) participants as a larger proportion of the population becomes infected or immune.[27]

### Benefits and risks of HICS in the context of epidemic suppression

A strategy of HICS followed by post-trial vaccination is superior to field trials in our model where R_eff_ is less than 1.14 (at 90% vaccine efficacy) to 1.3 (50% vaccine efficacy), both in terms of the reduction in population mortality from vaccination and the duration of transmission control measures (Figure 1). In this setting a HICS strategy has two additional advantages not considered in our simplified vaccine development timeline. First, HICS permit comparison of multiple candidate vaccines in the same population, free from phenomena that might compromise the interpretation of field trials including the interactions of indirect effects of multiple vaccines and the inclusion of non-comparable study populations in multi-centre trials.[12] Second, a HICS strategy permits testing vaccine efficacy despite very low rates of population transmission, and if suppression policies are maintained, additional delays (e.g., in vaccine development or population vaccination) do not erode potential benefits whereas this is a significant issue under mitigation policies (Figure A3, Supplementary Results).

Conducting a HICS in a setting of low background incidence is associated with higher marginal risk for participants (Figure 4), but this additional risk is very small, and the marginal risk does not meaningfully decrease across the range of values of R_eff_ where HICS are the preferred means of demonstrating vaccine efficacy (below 1.3). The ratio of population deaths avoided to deaths among HICS participants is at least 200 000 to one (and the likelihood of any death among HICS participants is very small).

### Potential exploitation of populations under mitigation or less stringent public health measures

Beyond practical caveats, there are also ethical questions about where field trails should be conducted.[12, 28] If field trials are conducted in higher incidence populations, where they provide the fastest results, the burdens of these trials would be concentrated where the burdens of disease are highest, but the benefits would largely accrue to other populations maintained under epidemic suppression. In some cases, this may be considered mutually beneficial (e.g., where populations have democratically chosen mitigation), but in other cases it may be considered problematic exploitation (e.g., where a majority of a population would prefer suppression but the local public health policy achieves little or no transmission control).

### Limitations

As with any modelling approach, the major limitations of our findings relate to the assumptions and inputs of the model. The assumptions with the greatest potential effect on our findings are the structural assumptions of a compartmental epidemiological model noted above, and the timeline presented in Figure A2 of the Supplementary Methods. This timeline includes a number of assumptions that are largely favourable to field trials (including the feasibility of recruitment and identification of mostly mild incident infection among a large sample of susceptible individuals over a prolonged trial duration[15]) and to the effectiveness of vaccination as a method of epidemic termination in general (including the time to the start of efficacy trials and the speed and reach of a population vaccination campaign). As noted in the sensitivity analysis, these delays have the greatest impact under mitigation (where field trials are preferred to HICS), and minimal effect under suppression (where HICS are preferred). Our assumption of a sequential HICS and expanded safety trial (as proposed elsewhere[4]) is the source of the disadvantage seen for HICS under a mitigation approach; running these components in parallel would abolish any speed or mortality advantage of field trials. We have also modelled a simple four-phase approach to transmission control measures, but more complex patterns of transmission control could be incorporated into the same methods should such an approach be formulated.

The IFR of COVID-19 (both the population average and the estimate for HICS participants) acts as a simple factor in the mortality effects of mitigation and vaccination and does not alter the relative conclusions we present. The inclusion of R_eff_ as a variable in all analyses allows readers with different estimates for R_0_ and the infectious period of COVID-19 to substitute these values for ours by calculating the infectious coefficient β.

Throughout our analysis we have assumed that regulators would be prepared to provide emergency use authorisation for a vaccine based on the results of a HICS and expanded safety trial.[22] Although concerns regarding generalisability (e.g., differential efficacy of vaccination between healthy HICS participants and the general population) are reasonable, such potential disadvantages would be counterbalanced by the opportunity to select the most effective vaccine, among a wide range of candidate vaccines in the HICS strategy, and through assessments of relative immunogenicity during an expanded safety trial. The vaccination efficacy variable in our model represents average efficacy in the general population so as to estimate population benefits accurately – a higher vaccine efficacy among HICS participants would only serve to reduce the required group size and the risk to which participants are exposed.

### Conclusions

The maximal benefit of vaccination against SARS-CoV-2 accrues to populations maintained under epidemic suppression. Within these populations, HICS are the only option able to demonstrate vaccine efficacy, and the marginal risks to HICS participants are extremely low. Field trials may be faster than HICS under a strategy of optimal mitigation, but such strategies invariably result in less vaccination benefit to the local population, and additional unforeseen delays or imperfect public health measures risk no benefit at all. This could lead to ethically problematic exploitation of higher incidence populations, whose participation in field trials primarily benefits other (lower incidence) populations.

## FOOTNOTE

All authors attest they meet the ICMJE criteria for authorship.

No conflicts of interest to declare (all authors).

This research did not receive any specific grant from funding agencies in the public, commercial, or not-for-profit sectors.

This work has not been presented previously.

## Data Availability

All input data and model specifications are provided in the manuscript.

## SUPPLEMENTARY METHODS

### Compartmental model structure

**Figure A1:**
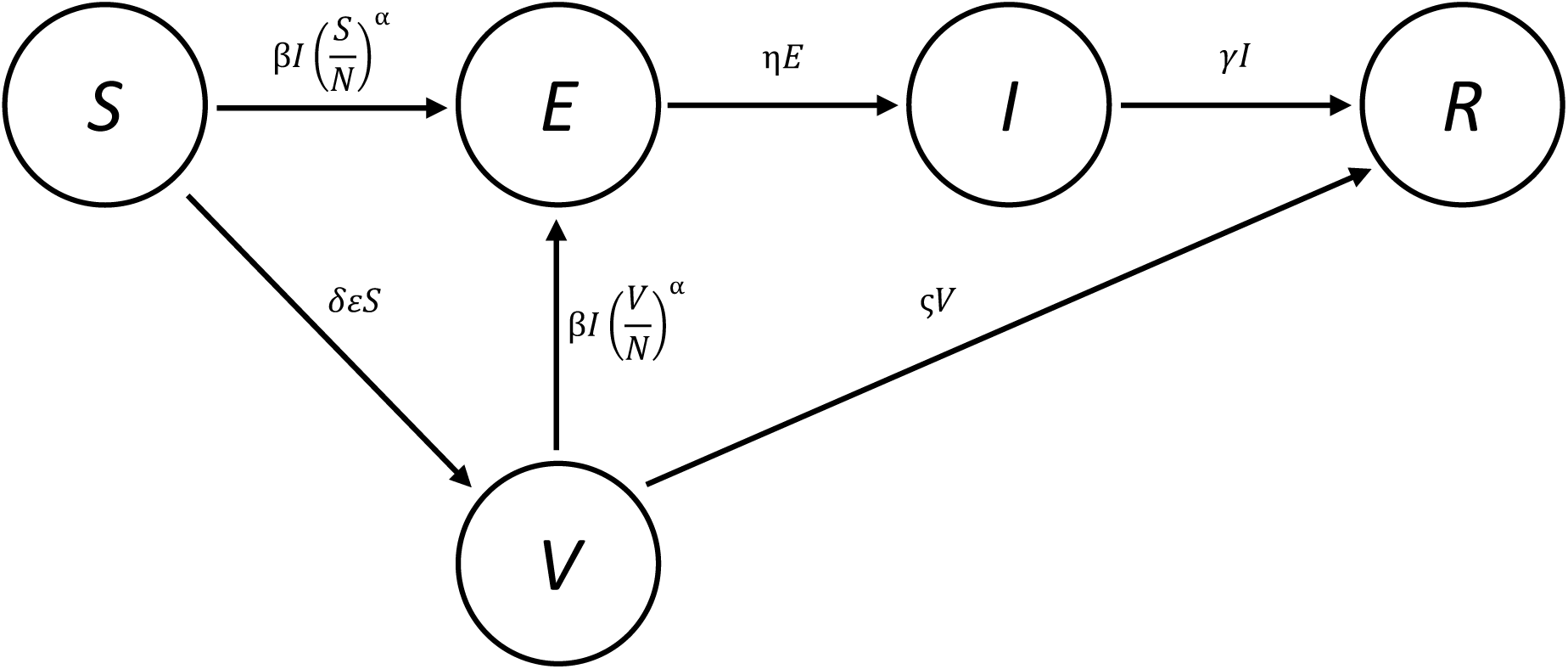
SEIRV model with transition forces

### Model equations

The flow of individuals through the compartments of the model is governed by a set of ordinary differential equations:

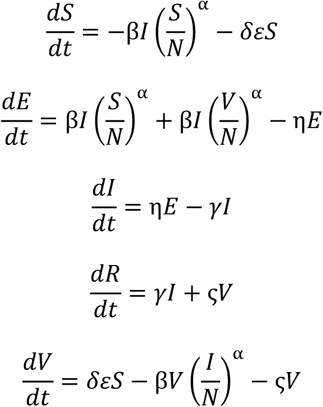

The daily incident mortality is calculated by:

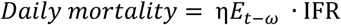

**Table A1:**
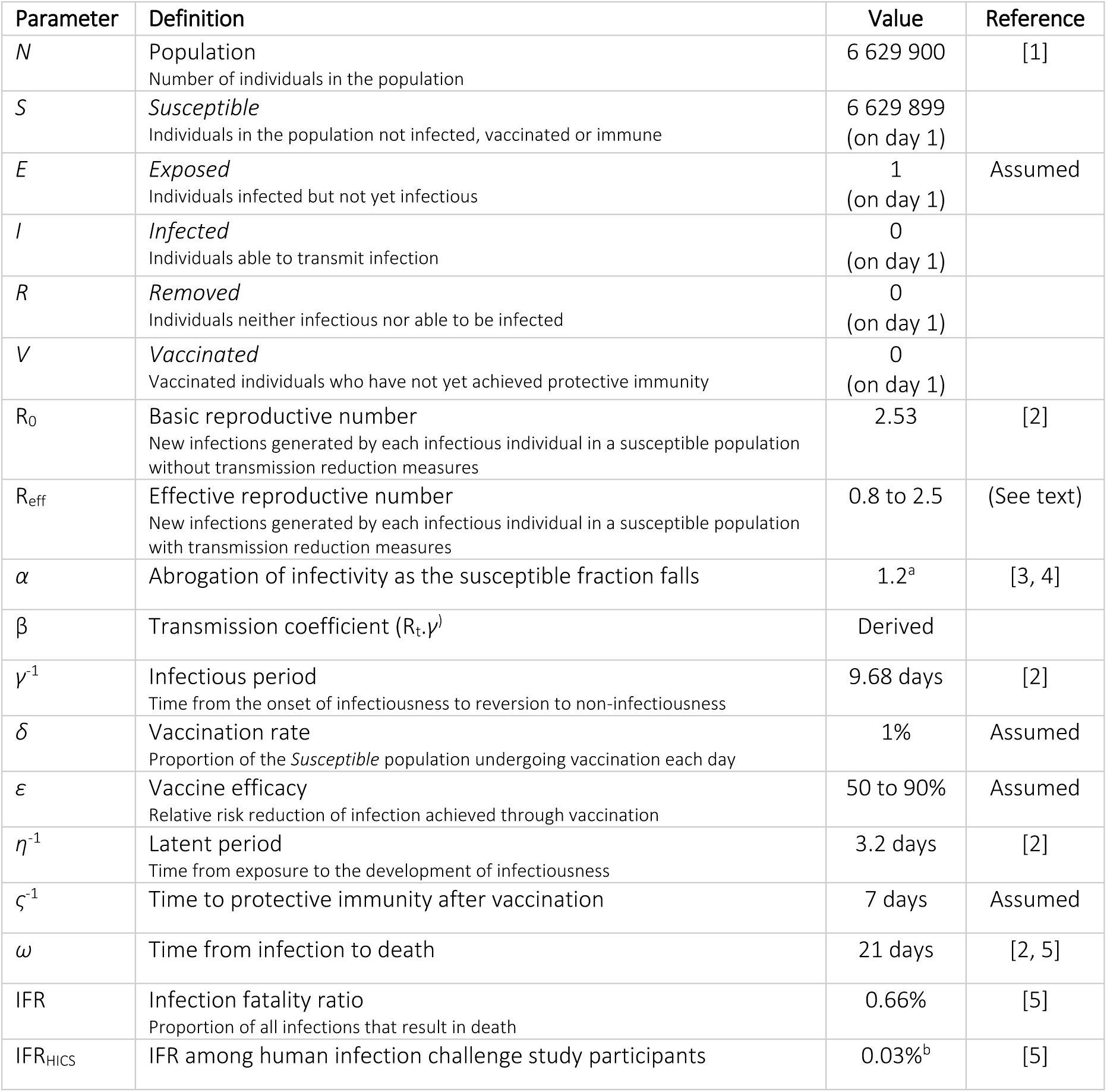
Model parameters

As a comparison, the same methods applied to publicly-available Italian data but omitting the time adjustment in view of the declining epidemic in Italy give an estimated IFR of 0.02% (29 858[6] cases among 9 567 000[7] individuals aged 50-59 years gives an estimated 19 050 symptomatic cases and 40 532 total infections among individuals aged 20-29 years, of whom 7 have died [6]).

Neither of these estimates account for absolute under-ascertainment of symptomatic cases, the significance of which remains uncertain in the absence of reliable data on population incidence. A recent assessment of the seasonally-adjusted excess mortality during the COVID-19 epidemic in Italy compared to previous years suggests an IFR of 0.006% among individuals aged 20-29 years in the context of an overall IFR of 0.84%.[8] This analysis assumed that all changes in mortality during the epidemic in Italy were due to COVID-19, and it is possible that a coincidental reduction in death from trauma or other non-infectious causes artificially depresses this estimate.

We also anticipate that HICS participants would be free from significant comorbidities, and perhaps have an even lower IFR than their age-matched peers. The marked over-representation of young people with comorbidities among COVID-19 deaths in New York City[9] (80% in what is presumably a predominantly disease-free subpopulation) suggests that healthy volunteers may have a substantially lower IFR than suggested by their age alone.

Overall, we believe that the estimate of IFR in challenge study participants used in our analysis (0.03%) is pessimistic and likely over-states the risk to these individuals.

**Figure A2.**
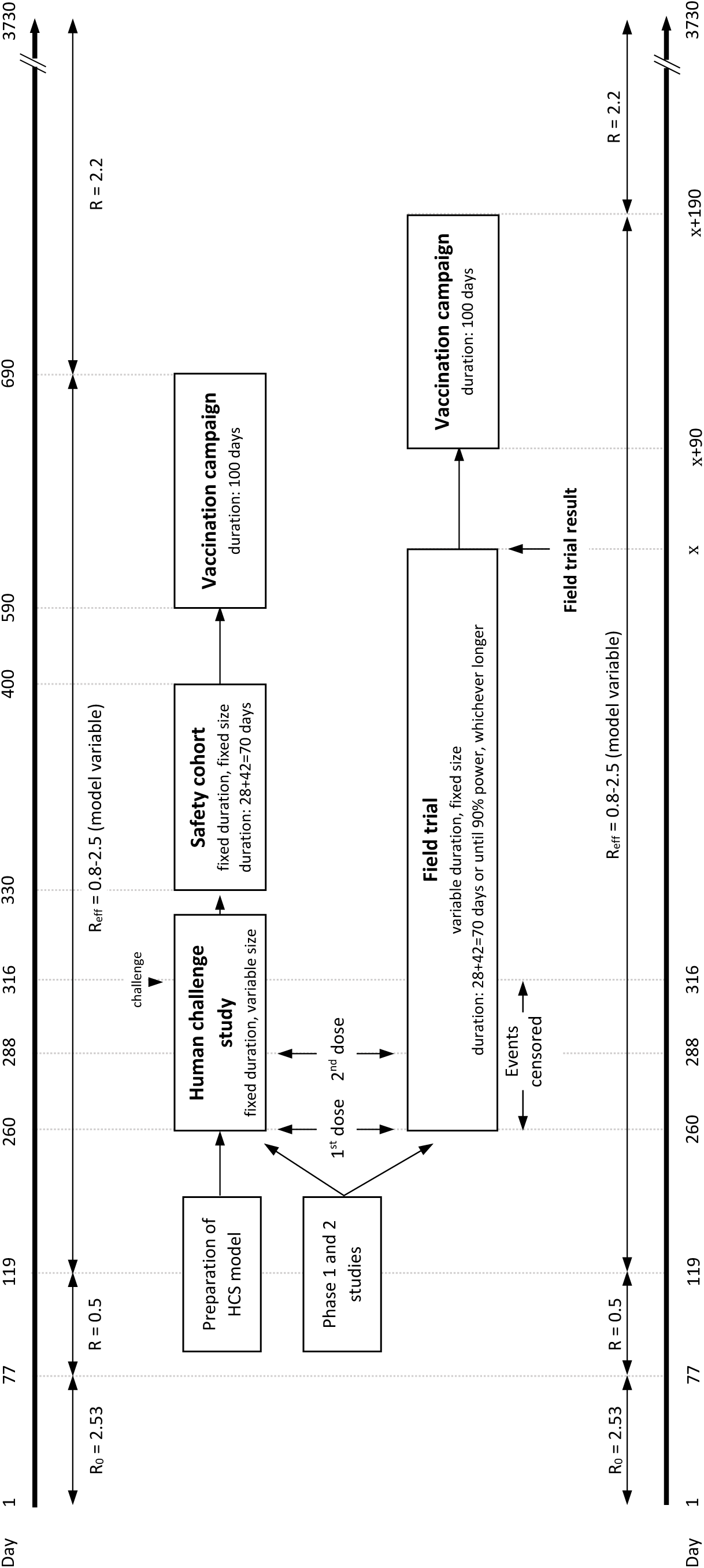
Modelled timeline

### Power calculations

The group size for the two randomised controlled trials is given by the following standard formula:

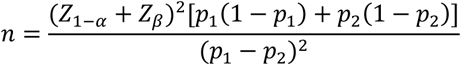

Where: *Z* is the cumulative distribution function of a standardised normal deviate

*α* is the one-tailed type 1 error rate (set at 2.5%)

β is the type 2 error rate

*p*_1_ is the probability of infection in the placebo group × (1- minimum vaccine efficacy

*p*_2_ is the probability of infection in the vaccinated group

For the challenge study, the equation is solved for *n* using the desired statistical power and the proportion of participants with successful experimental infection (90% of the placebo group).

For the field trial, this equation is solved for β (statistical power) using the proportion of the population with incident infection after the initial censoring interval (*p*_1_) and the (smaller) number of participants in the placebo group who remain uninfected between day -56 (recruitment) and day 1 (*n*). This per protocol analysis was chosen based on the inherent assumption of equal attack rate across the population in the compartmental model (i.e., no post-randomisation selection bias), to match the study question addressed by the challenge study (vaccine efficacy in fully-vaccinated uninfected individuals), and to address anticipated regulatory requirements for demonstration of non-situational vaccine efficacy[10].

Table A2 presents an example calculation drawn from the model, where R_eff_ is set to 1.2 and vaccine efficacy to 70%. The model used for power calculations does not include a lifting of transmission control measures at three years to allow calculation of the maximum trial duration.

**Table A2:**
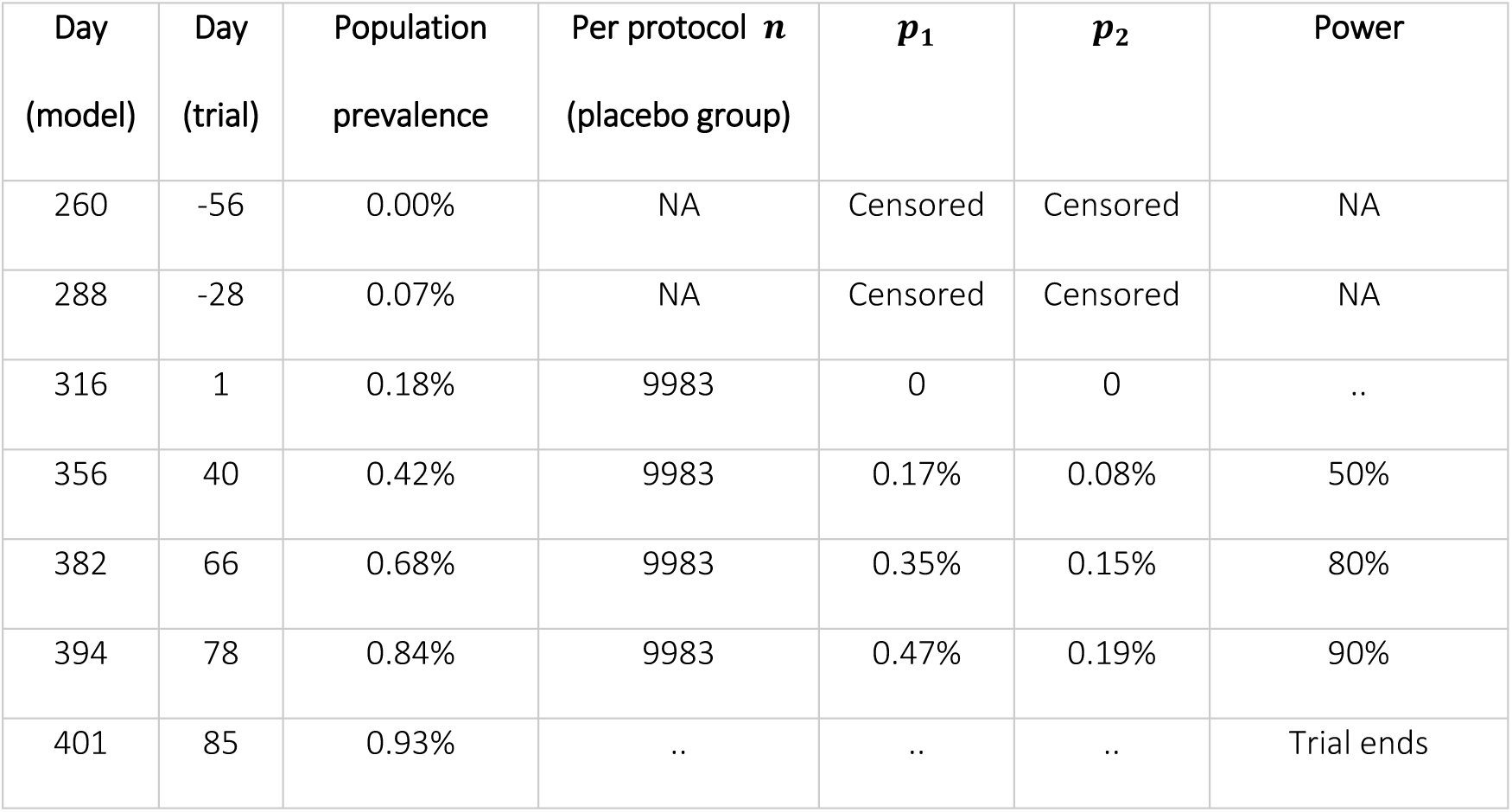
Example field trial power calculation

| Day<br>(model) | Day<br>(trial) | Population<br>prevalence | Per protocol $n$<br>(placebo group) | $p_1$ | $p_2$ | Power |
| --- | --- | --- | --- | --- | --- | --- |
| 260 | -56 | 0.00% | NA | Censored | Censored | NA |
| 288 | -28 | 0.07% | NA | Censored | Censored | NA |
| 316 | 1 | 0.18% | 9983 | 0 | 0 | .. |
| 356 | 40 | 0.42% | 9983 | 0.17% | 0.08% | 50% |
| 382 | 66 | 0.68% | 9983 | 0.35% | 0.15% | 80% |
| 394 | 78 | 0.84% | 9983 | 0.47% | 0.19% | 90% |
| 401 | 85 | 0.93% | .. | .. | .. | Trial ends |

## SUPPLEMENTARY RESULTS

**Figure A3.**
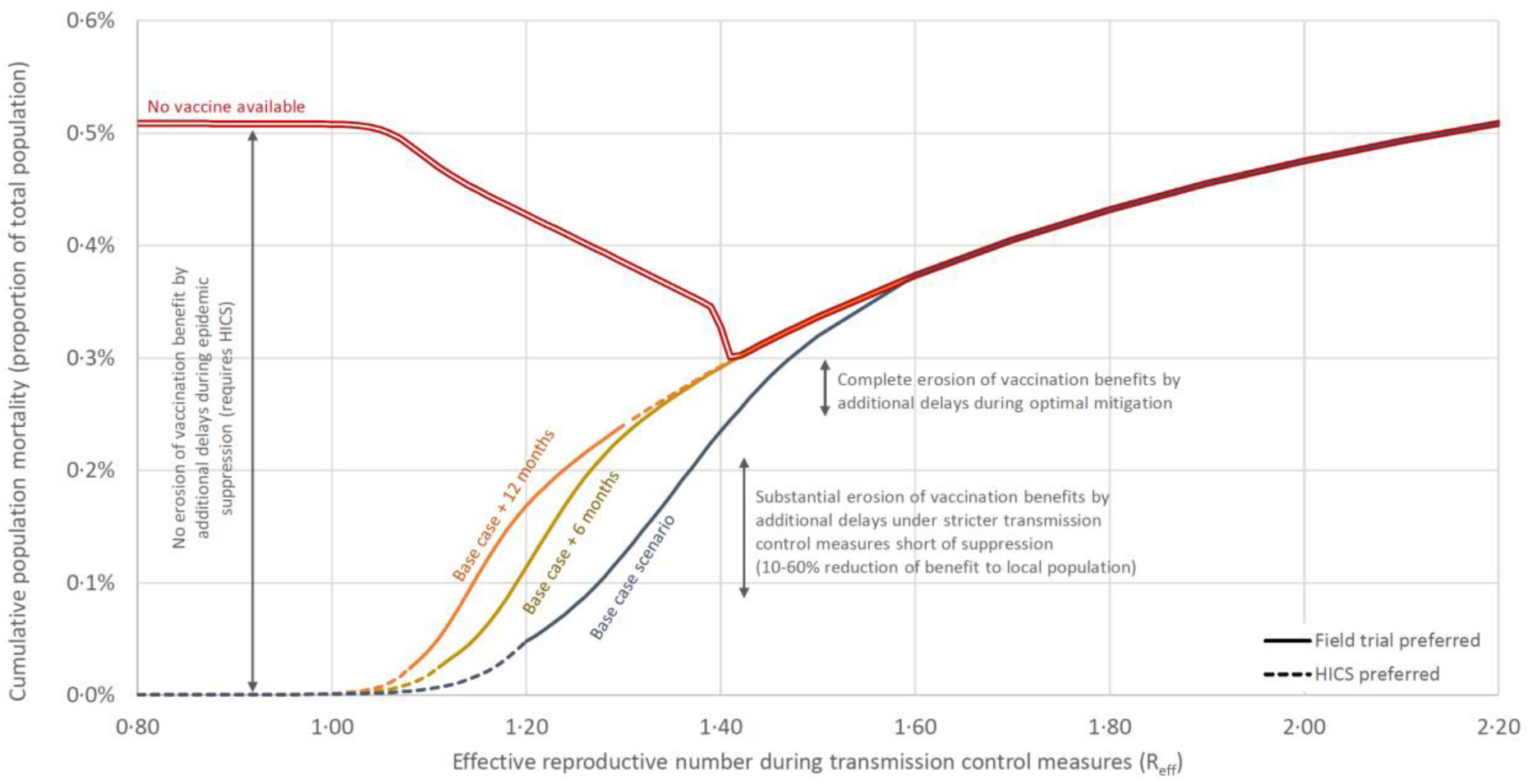
Effect of additional non-trial delays to vaccine development timeline on the benefit of post-trial vaccination to the local population (70% vaccine efficacy).

The delays presented in Figure A3 include the time from transmission control measures to the start of efficacy trials (183 days in the base case scenario presented in the manuscript), the delay from the end of an efficacy trial to the start of a population vaccination campaign (90 days), and the time to complete vaccination coverage in the population (100 days). These delays are generally additive, although the rate of population vaccination has more complex effects on epidemic termination than the two other delays. Any additional trial duration required for vaccine safety outcomes above the 70 days included the base case scenario could also be considered as an additive delay.

As seen in the figure, additional delays of 12 months (or longer) have minimal effect on the benefit of vaccination to the local population under conditions of epidemic suppression, because the rate of accumulation of new cases (and infection-related mortality) is very slow. Under these conditions, a human infection challenge study (HICS) is the only mechanism by which vaccine efficacy can be demonstrated within the local population. In contrast, under conditions of epidemic mitigation, the benefit of population vaccination is rapidly eroded with additional delays. Under optimal mitigation (R_eff_ 1.41), no benefit accrues to the local population in the setting of a six-month additional delay to the vaccine development timeline.

**Figure A4.**
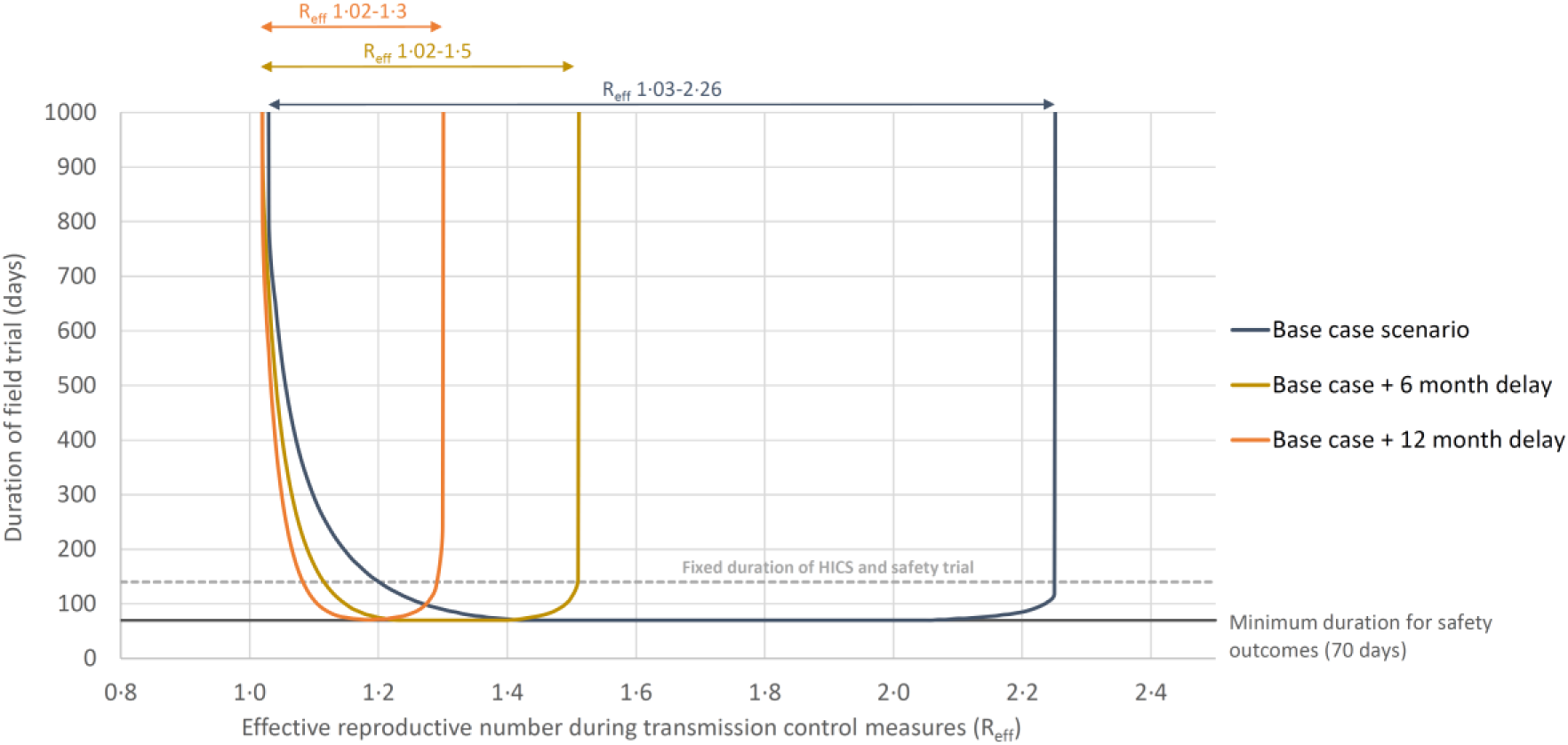
Effect of additional non-trial delays on the feasibility of vaccine field trial (70% vaccine efficacy).

These delays also have an important effect on the feasibility of vaccine field trials (achievement of 90% power to demonstrate vaccine efficacy greater than 30% with one-sided alpha 2.5%). As seen in Figure A4, additional delays to the start of a field trial beyond the 183 days (the base case scenario) rapidly restrict the range of R_eff_ over which such a trial is feasible (but not necessarily preferred to HICS or beneficial to the local population). An additional 6 month delay to the start of a field trial reduces this feasibility window to values of R_eff_ 1.02-1.5; an additional 12 month delay reduces the window to R_eff_ 1.02-1.3.

**Figure A5.**
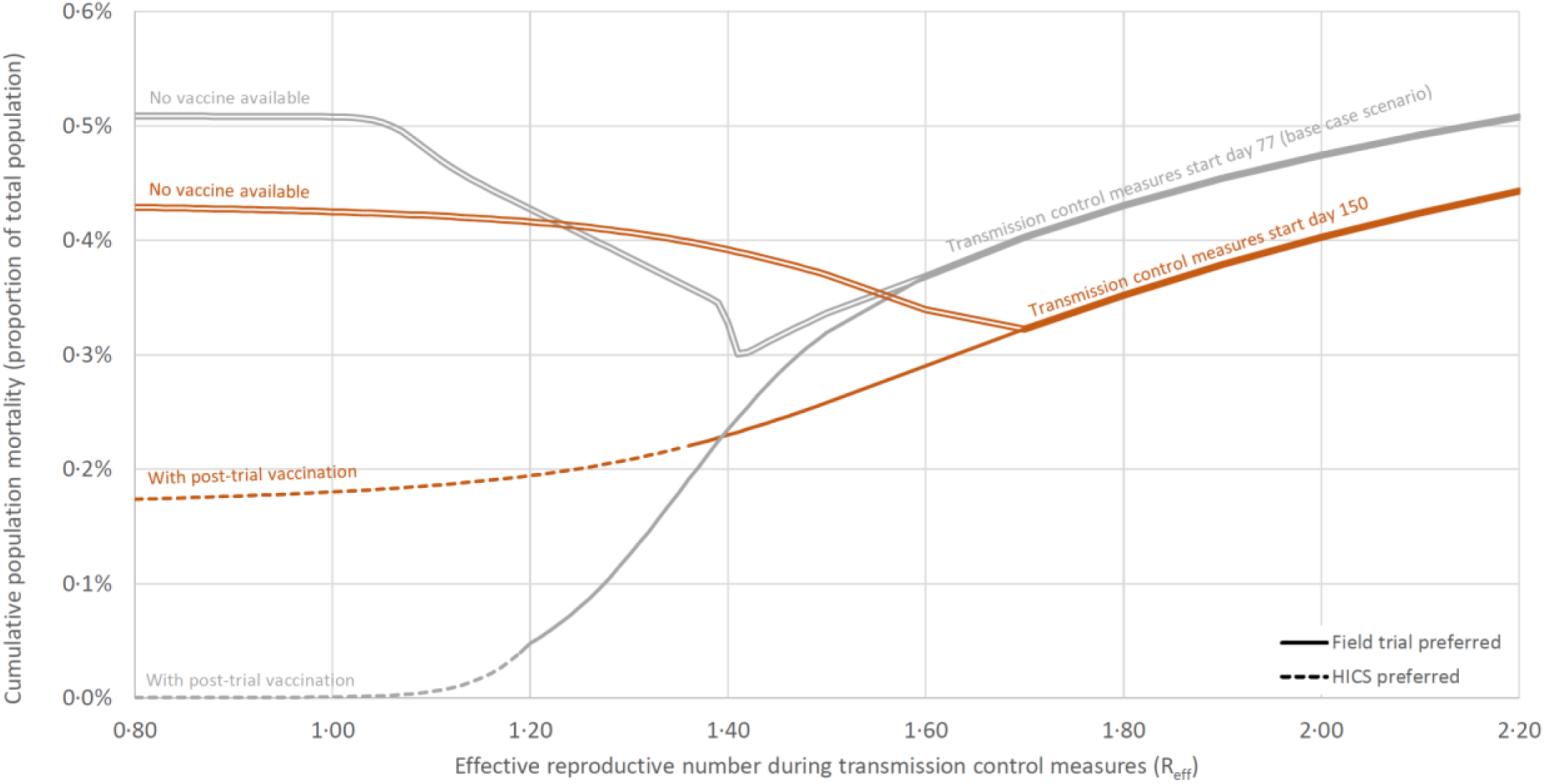
Effect of delayed institution of transmission control measures on the feasibility of vaccine trials and benefit of post-trial vaccination (70% vaccine efficacy).

Figure A5 shows the effect of the delayed institution of transmission control measures beyond day 77 (the day of the first expected death in the uncontrolled initial phase of the model). The four-phase pattern of uncontrolled transmission, a brief period of tight transmission control measures, a partial relaxation, then a return to normal conditions is not dissimilar to the planned response to COVID-19 in a number of different countries.

In this figure, the base case scenario is compared to the institution of tight transmission control measures on day 150, resulting in a peak pre-control population mortality rate of 2.3 deaths per 100,000 population per day[9] and a cumulative population prevalence of infection of 0.91% assuming 0.66% IFR. The start of the vaccine efficacy trials is fixed at 183 days after the introduction of transmission control measures, as in the base case scenario.

As seen in the figure, a higher proportion of *Removed* individuals resulting from more pre-control infections both decreases the maximal benefit of vaccination and increases the range of R_eff_ that permits some residual benefit. The relative benefit of field trials and HICS remains unchanged - higher transmission rates and optimal mitigation favours field trials, but stricter control offers greater vaccination benefits dependent on HICS.

